# De novo missense variants in *SLC32A1* cause a neurodevelopmental disorder with epilepsy due to impaired GABAergic neurotransmission

**DOI:** 10.1101/2021.12.06.21267233

**Authors:** Konrad Platzer, Heinrich Sticht, Caleb Bupp, Mythily Ganapathi, Elaine M. Pereira, Gwenaël Le Guyader, Frederic Bilan, Lindsay B. Henderson, Holger Taschenberger, Nils Brose, Rami Abou Jamra, Sonja M. Wojcik

## Abstract

We describe four patients with a neurodevelopmental disorder and *de novo* missense variants in *SLC32A1,* the gene that encodes the vesicular GABA transporter (VGAT). The main phenotype comprises moderate to severe intellectual disability, early onset epilepsy within the first 18 months of life and a choreatic, dystonic or dyskinetic movement disorder. *In silico* modeling and functional analyses in cultured neurons reveal that three of these variants, which are located in helices that line the putative GABA transport pathway, result in reduced quantal size, consistent with impaired filling of synaptic vesicles with GABA. The fourth variant, located in the VGAT N-terminus, does not affect quantal size, but increases presynaptic release probability, leading to more severe synaptic depression during high frequency stimulation. Thus, variants in VGAT can impair GABAergic neurotransmission via at least two mechanisms, by affecting synaptic vesicle filling and by altering synaptic short-term plasticity. This work establishes *de novo* missense variants in *SLC32A1* as a novel cause for a neurodevelopmental disorder with epilepsy.

## Introduction

Members of the solute carrier (SLC) protein family transport a wide range of substrates with amino acids being the most frequent.^1^ The SLC family 32 Member 1 (SLC32A1) is the only protein identified to date, that loads gamma-aminobutyric acid (GABA) and glycine into synaptic vesicles (SVs), and is therefore also known as the vesicular GABA transporter (VGAT) or vesicular inhibitory amino acid transporter (VIAAT). While GABA is an important inhibitory neurotransmitter crucial for the proper function of the mature brain, both GABA and glycine act as inhibitory neurotransmitters in the spinal cord and brainstem.

Homozygous *Slc32a1* knockout mice (*Slc32a1^KO^*) are embryonically lethal^2^, while heterozygotes show no abnormalities in behavioral assays.^3^ As disruption of GABAergic neurotransmission is an established cause of epilepsy and neurodevelopmental disorders^4^, impaired VGAT function constitutes a plausible cause for a neurodevelopmental disorder with epilepsy. Rare heterozygous missense variants in *SLC32A1* have recently been described to segregate in families with a phenotype of genetic epilepsy with febrile seizures plus (GEFS+) and idiopathic generalized epilepsy (IGE)^5^. Affected family members showed a wide range of seizure types, e.g. with febrile seizures, focal seizures, generalized seizures and unclassified seizures, but no developmental delay (DD) or intellectual disability (ID).

In this study, we report on four individuals with *de novo* missense variants in *SLC32A1* and a neurodevelopmental disorder with early onset epilepsy. We establish causality of the variants via *in silico* modelling and their functional evaluation in a murine neuronal cell culture model.

## Material and Methods

### Study subjects

This study was approved by the ethics committee of the University of Leipzig (402/16-ek). With the help of GeneMatcher^6^ we identified four individuals harboring heterozygous *de novo* missense variants in *SLC32A1* with an overlapping phenotype of a neurodevelopmental disorder with DD/ID and epilepsy at different centers in Germany, France and the USA. The collaborating physicians provided detailed clinical information via a uniform clinical questionnaire. Informed consent was obtained from all examined individuals or their legal guardians. For some cases, testing was done as part of routine clinical care, and therefore institutional ethics approval was not required. If done in a research setting, the testing was approved by the ethics committees of the respective centers. All families provided informed consent for clinical testing and publication.

### Identification and evaluation of variants

Exome sequencing using standard commercial products within diagnostic or research settings was performed in all four families. Since no causative variant was identified in a known disease gene in any of the four index individuals, evaluation of the exome sequencing data was performed to potentially identify variants in candidate genes. To prioritize variants in known disease as well as candidate genes, the following aspects were considered: gene and variant attributes, a minor allele frequency of below 1 % in the general population, assumed effect on protein function, *in silico* prediction tools, phenotype, family history, and inheritance according to the local proceedings in the respective centers. *De novo* occurrence of the variants in *SLC32A1* was confirmed in all individuals. For *in silico* prediction of missense variants, the following tools were used: CADD, REVEL, MutationTaster, M-CAP, PolyPhen-2, GERP++^7–12^. Families of individuals 1-3 underwent trio exome sequencing.

Clinical trio exome sequencing was performed for individual 3 as previously described^13^ and subsequently confirmed with Sanger sequencing. The *de novo* variant in individual 4 was identified by singleton exome sequencing and subsequent validation of *de novo* occurrence with Sanger sequencing. The databases of the Genome Aggregation Database (gnomAD, v2.1.1) served as control populations.^14^

### Structural modelling

The structure of VGAT was modelled with HHpred^15^ and Modeller^16^ using the crystal structure of the transporter SLC38A9 (PDB: 6C08;^17^) as a template. The arginine substrate present in the template structure was replaced by GABA. The effect of the sequence variants was investigated with the tools VIPUR^18^ and Missense3D.^19^ VMD^20^ and RasMol^21^ were used for structure analysis and visualization.

### Lentiviral constructs

Murine VGAT cDNA was amplified from a C57Bl/6N cDNA library using the primers 5’-CGGCTAGCGCGAGGGTCATGAGCCAGAGC-3’ and 5’-GCTTCGAATGGGGGTGGGGGCGGGAATGAC-3’. PCR products were subcloned into the pCR^TM^II-TOPO vector (ThermoFisher). Each variant was then introduced using QuikChange mutagenesis (Agilent). WT and the four mutated VGAT cDNAs were then subcloned using the primer derived NheI and BstBI sites into a lentiviral vector, which expresses the target gene under control of a Synapsin-1 promoter and an EGFP reporter from a separate ubiquitin C romoter.^22^ Lentiviral particles were produced according to published protocols.^23, 24^

### Neuron culture and electrophysiological recordings

Autaptic microisland cultures of striatal neurons were prepared from E17.5 mouse embryos as previously described.^2, 25, 26^ Cultures were infected with lentiviral particles on day in vitro (DIV) 2-3 and patch-clamp recordings were performed between DIV 9-14. Whole-cell voltage clamp recordings were acquired at room temperature (22-25°C) using an EPC-10 amplifier and Patchmaster software (HEKA). Whole-cell capacitance was estimated from capacitive current transients in response to 10 mV step depolarizations assuming a simple two-compartment model.^27^ Capacitance values reported in Table 2 represent total cell capacitance, i.e. the sum of C_1_ (representing soma and proximal dendrites) and C_2_ (representing distal dendrites) of the two-compartment equivalent circuit model of Mennerick et al. (1995).^27^ The intracellular solution contained (in mM) 100 KCl, 40 K-Gluconate, 10 HEPES, 0.1 EGTA, 4 MgATP, 0.3 Na_2_GTP, 15 creatine phosphate, and 5 U/ml phosphocreatine kinase (315 mOsm/l, pH 7.2). The standard extracellular solution contained (in mM) 140 NaCl, 2.4 KCl, 10 HEPES, 10 glucose, 2 CaCl_2_, and 4 MgCl_2_ (305-307 mOsm/l, pH 7.4). For estimating I_sucrose_ elicited by hyperosmotic stimulation, the standard bath solution was supplemented with 500 mM sucrose and CaCl_2_ was omitted. A custom made fast-flow perfusion system consisting of glass capillaries controlled by a stepper motor, which allows a complete solution exchange around the recorded neuron, was used to rapidly switch between extracellular solutions. Inhibitory postsynaptic currents (IPSCs) were evoked by depolarizing voltage steps of 1 ms duration from the holding potential of −70 mV to −20 mV. Liquid junction potentials were not corrected for. The GABAergic nature of postsynaptic currents was confirmed by their sensitivity to 20 µM (-)-Bicuculline methiodide (Hello Bio). Single action potential (AP)-evoked IPSCs were recorded at an inter-stimulus interval of 10 s which was sufficiently long to allow for full recovery from synaptic depression, and 10-15 IPSCs were averaged for analysis. Trains of IPSCs consisting of 30 stimuli and evoked at stimulus frequencies of 10 Hz and 40 Hz were recorded three times for each cell, with 1.5 minute intervals in between successive trials, and averaged prior to analysis. Since miniature inhibitory postsynaptic current (mIPSC) frequency was found to be low in resting autaptic striatal neurons, mIPSCs were recorded after each 10 Hz train for 1.5 minutes, for a total duration of 4.5 minutes. For mIPSC recordings under conditions of elevated osmolarity (355-357 mOsm/l), 50 mM sucrose were added to the standard extracellular solution.

**Table 2.**
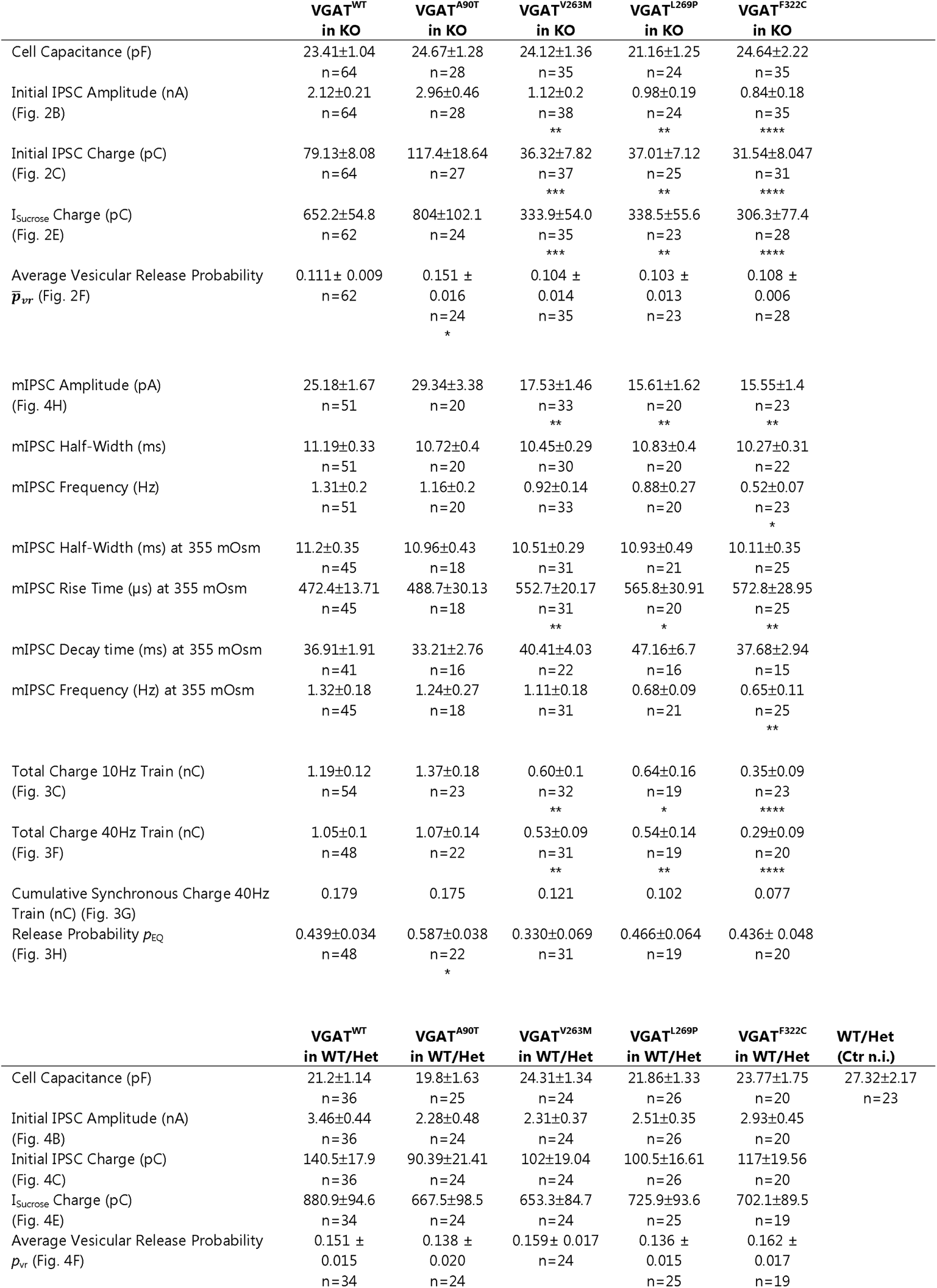
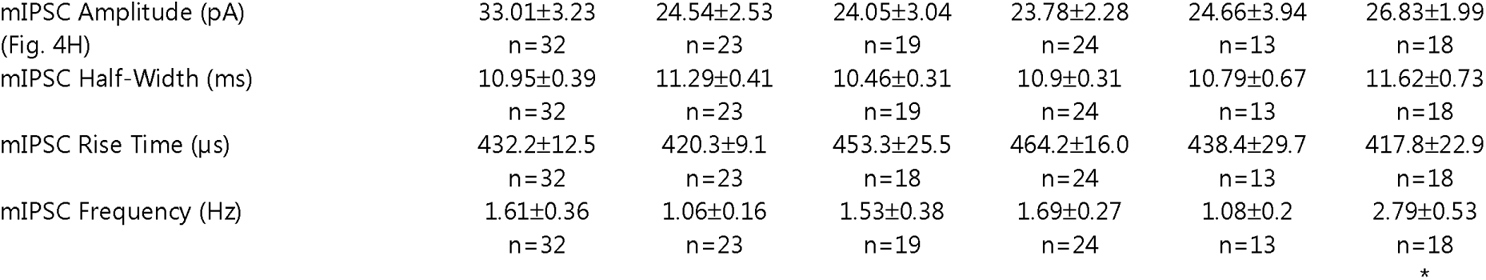
Characteristics of GABAergic synaptic transmission.

|  | VGAT<sup>WT</sup><br>in KO | VGAT<sup>A90T</sup><br>in KO | VGAT<sup>V263M</sup><br>in KO | VGAT<sup>L269P</sup><br>in KO | VGAT<sup>F322C</sup><br>in KO |  |
| --- | --- | --- | --- | --- | --- | --- |
| Cell Capacitance (pF) | 23.41±1.04<br>n=64 | 24.67±1.28<br>n=28 | 24.12±1.36<br>n=35 | 21.16±1.25<br>n=24 | 24.64±2.22<br>n=35 |  |
| Initial IPSC Amplitude (nA)<br>(Fig. 2B) | 2.12±0.21<br>n=64 | 2.96±0.46<br>n=28 | 1.12±0.2<br>n=38<br>** | 0.98±0.19<br>n=24<br>** | 0.84±0.18<br>n=35<br>**** |  |
| Initial IPSC Charge (pC)<br>(Fig. 2C) | 79.13±8.08<br>n=64 | 117.4±18.64<br>n=27 | 36.32±7.82<br>n=37<br>*** | 37.01±7.12<br>n=25<br>** | 31.54±8.047<br>n=31<br>**** |  |
| I<sub>Sucrose</sub> Charge (pC)<br>(Fig. 2E) | 652.2±54.8<br>n=62 | 804±102.1<br>n=24 | 333.9±54.0<br>n=35<br>*** | 338.5±55.6<br>n=23<br>** | 306.3±77.4<br>n=28<br>**** |  |
| Average Vesicular Release Probability<br>$\bar{p}_{vr}$ (Fig. 2F) | 0.111± 0.009<br>n=62 | 0.151 ±<br>0.016<br>n=24<br>* | 0.104 ±<br>0.014<br>n=35 | 0.103 ±<br>0.013<br>n=23 | 0.108 ±<br>0.006<br>n=28 | |
| mIPSC Amplitude (pA)<br>(Fig. 4H) | 25.18±1.67<br>n=51 | 29.34±3.38<br>n=20 | 17.53±1.46<br>n=33<br>** | 15.61±1.62<br>n=20<br>** | 15.55±1.4<br>n=23<br>** |  |
| mIPSC Half-Width (ms) | 11.19±0.33<br>n=51 | 10.72±0.4<br>n=20 | 10.45±0.29<br>n=30 | 10.83±0.4<br>n=20 | 10.27±0.31<br>n=22 |  |
| mIPSC Frequency (Hz) | 1.31±0.2<br>n=51 | 1.16±0.2<br>n=20 | 0.92±0.14<br>n=33 | 0.88±0.27<br>n=20 | 0.52±0.07<br>n=23<br>* |  |
| mIPSC Half-Width (ms) at 355 mOsm | 11.2±0.35<br>n=45 | 10.96±0.43<br>n=18 | 10.51±0.29<br>n=31 | 10.93±0.49<br>n=21 | 10.11±0.35<br>n=25 |  |
| mIPSC Rise Time (μs) at 355 mOsm | 472.4±13.71<br>n=45 | 488.7±30.13<br>n=18 | 552.7±20.17<br>n=31<br>** | 565.8±30.91<br>n=20<br>* | 572.8±28.95<br>n=25<br>** |  |
| mIPSC Decay time (ms) at 355 mOsm | 36.91±1.91<br>n=41 | 33.21±2.76<br>n=16 | 40.41±4.03<br>n=22 | 47.16±6.7<br>n=16 | 37.68±2.94<br>n=15 |  |
| mIPSC Frequency (Hz) at 355 mOsm | 1.32±0.18<br>n=45 | 1.24±0.27<br>n=18 | 1.11±0.18<br>n=31 | 0.68±0.09<br>n=21 | 0.65±0.11<br>n=25<br>** |  |
| Total Charge 10Hz Train (nC)<br>(Fig. 3C) | 1.19±0.12<br>n=54 | 1.37±0.18<br>n=23 | 0.60±0.1<br>n=32<br>** | 0.64±0.16<br>n=19<br>* | 0.35±0.09<br>n=23<br>**** |  |
| Total Charge 40Hz Train (nC)<br>(Fig. 3F) | 1.05±0.1<br>n=48 | 1.07±0.14<br>n=22 | 0.53±0.09<br>n=31<br>** | 0.54±0.14<br>n=19<br>** | 0.29±0.09<br>n=20<br>**** |  |
| Cumulative Synchronous Charge 40Hz<br>Train (nC) (Fig. 3G) | 0.179 | 0.175 | 0.121 | 0.102 | 0.077 |  |
| Release Probability $p_{EQ}$<br>(Fig. 3H) | 0.439±0.034<br>n=48 | 0.587±0.038<br>n=22<br>* | 0.330±0.069<br>n=31 | 0.466±0.064<br>n=19 | 0.436± 0.048<br>n=20 | |
|  | VGAT<sup>WT</sup><br>in WT/Het | VGAT<sup>A90T</sup><br>in WT/Het | VGAT<sup>V263M</sup><br>in WT/Het | VGAT<sup>L269P</sup><br>in WT/Het | VGAT<sup>F322C</sup><br>in WT/Het | WT/Het<br>(Ctr n.i.) |
| Cell Capacitance (pF) | 21.2±1.14<br>n=36 | 19.8±1.63<br>n=25 | 24.31±1.34<br>n=24 | 21.86±1.33<br>n=26 | 23.77±1.75<br>n=20 | 27.32±2.17<br>n=23 |
| Initial IPSC Amplitude (nA)<br>(Fig. 4B) | 3.46±0.44<br>n=36 | 2.28±0.48<br>n=24 | 2.31±0.37<br>n=24 | 2.51±0.35<br>n=26 | 2.93±0.45<br>n=20 |  |
| Initial IPSC Charge (pC)<br>(Fig. 4C) | 140.5±17.9<br>n=36 | 90.39±21.41<br>n=24 | 102±19.04<br>n=24 | 100.5±16.61<br>n=26 | 117±19.56<br>n=20 |  |
| I<sub>Sucrose</sub> Charge (pC)<br>(Fig. 4E) | 880.9±94.6<br>n=34 | 667.5±98.5<br>n=24 | 653.3±84.7<br>n=24 | 725.9±93.6<br>n=25 | 702.1±89.5<br>n=19 |  |
| Average Vesicular Release Probability<br>$p_{vr}$ (Fig. 4F) | 0.151 ±<br>0.015<br>n=34 | 0.138 ±<br>0.020<br>n=24 | 0.159± 0.017<br>n=24 | 0.136 ±<br>0.015<br>n=25 | 0.162 ±<br>0.017<br>n=19 | |
| mIPSC Amplitude (pA)<br>(Fig. 4H) | 33.01±3.23<br>n=32 | 24.54±2.53<br>n=23 | 24.05±3.04<br>n=19 | 23.78±2.28<br>n=24 | 24.66±3.94<br>n=13 | 26.83±1.99<br>n=18 |
| mIPSC Half-Width (ms) | 10.95±0.39<br>n=32 | 11.29±0.41<br>n=23 | 10.46±0.31<br>n=19 | 10.9±0.31<br>n=24 | 10.79±0.67<br>n=13 | 11.62±0.73<br>n=18 |
| mIPSC Rise Time (μs) | 432.2±12.5<br>n=32 | 420.3±9.1<br>n=23 | 453.3±25.5<br>n=18 | 464.2±16.0<br>n=24 | 438.4±29.7<br>n=13 | 417.8±22.9<br>n=18 |
| mIPSC Frequency (Hz) | 1.61±0.36<br>n=32 | 1.06±0.16<br>n=23 | 1.53±0.38<br>n=19 | 1.69±0.27<br>n=24 | 1.08±0.2<br>n=13 | 2.79±0.53<br>n=18<br>* |

Spontaneously occurring mIPSCs were detected in continuous recordings by a template matching algorithm^28^ implemented in Igor Pro (Wavemetrics). Analyses were performed using Igor Pro and Prism (GraphPad).

### Western Blot

Western blotting was done on striatal mass cultures which were prepared according to the same protocols as autaptic cultures,^2, 25^ but plated on poly-L-lysine at a density of 7000 cells/cm^2^ and kept in Dulbecco’s modified eagle’s medium with GlutaMAX^TM^ (GIBCO, 31966-021) with 10% fetal bovine serum and B27-supplement (Gibco, 17504-044) for DIV 1. Cultures were infected on DIV 2 and harvested on DIV 9. Antibodies used for Western blotting were mouse-anti-Synapsin (Synaptic Systems, 106011) at 1:6000, rabbit-anti-VGAT (Synaptic Systems, 131013) at 1:1000 and mouse-anti-EGFP (Roche, 11814460001) at 1:3000.

### Statistics

Electrophysiological data are presented as mean ± standard error of the mean (SEM). Statistical significance was assessed using Kruskal-Wallis with post-hoc Dunn’s correction for multiple comparisons. To obtain estimates for the mean vesicular release probability 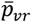 from IPSC vs I_sucrose_ scatter plots (Fig. 2F, Fig. 4F, Table 2), the matrix consisting of IPSC^i,gt^ and I_sucrose_^i,gt^, where i and gt denote cell index and genotype, respectively, was fitted with a linear model (intercept forced to zero) using R (R Foundation for Statistical Computing) with I_sucrose_ being the predictor variable and gt the interaction term, and statistical significance values (p-value) were adjusted for multiple comparisons using Tukey’s method (R package ‘emmeans’). SEM estimates for release probabilities *p*_EQ_ derived from line fits to Elmqvist-Quastel plots (Fig. 3H) were obtained by bootstrap resampling analysis using a balanced bootstrap approach implemented in Igor Pro (every experimental observation appeared exactly the same number of times in the total population of 2,000 bootstrap samples),^29, 30^ and statistical significance values (p-value) were adjusted for multiple comparisons using Bonferroni’s method.

**Figure 2:**
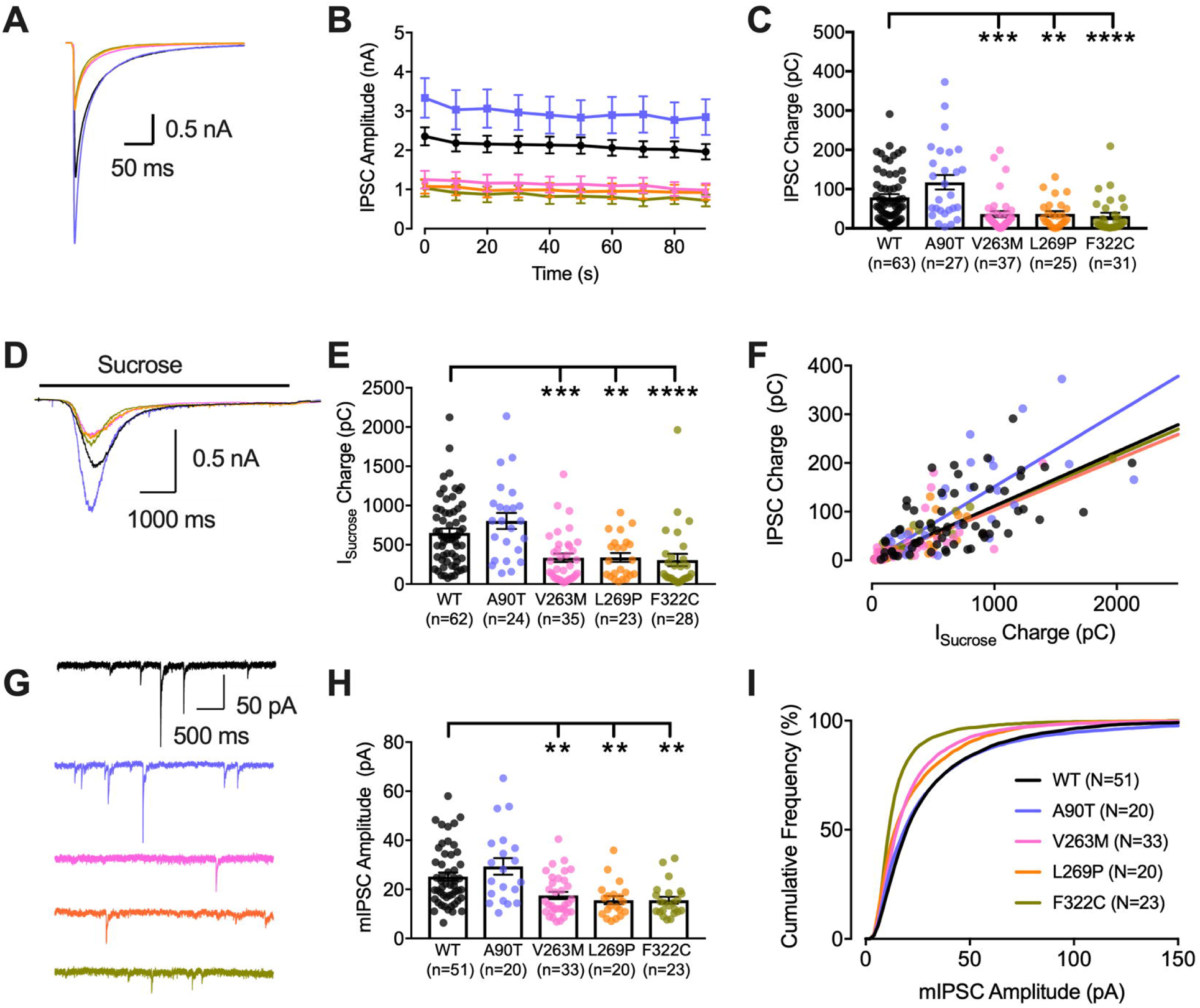
GABAergic IPSCs and mIPSCs in striatal neurons. (A) Example traces of single evoked inhibitory postsynaptic currents (IPSCs) from autaptic striatal *Slc32a1*^KO^ neurons with lentiviral expression of VGAT^WT^ (black), VGAT^A90T^ (blue), VGAT^V263M^ (pink), VGAT^L269P^ (orange), or VGAT^F322C^ (green). (B) VGAT^V263M^, VGAT^L269P^ and VGAT^F322C^ expressing neurons stimulated at 10 s intervals show reduced IPSC amplitude and (C) reduced IPSC charge. (D) Averaged traces during the application of 0.5 M sucrose which releases all fusion-competent SVs. (E) Reduced I_sucrose_ in neurons expressing VGAT^V263M^, VGAT^L269P^ or VGAT^F322C^. (F) Average vesicular release probability 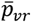, obtained by linear regression analysis of IPSC charge vs. I_sucrose_ charge, is higher for VGAT^A90T^. (G) Example traces of mIPSCs, representing the release of single SVs. (H) Average mIPSC amplitudes are significantly reduced in neurons expressing VGAT^V263M^, VGAT^L269P^ or VGAT^F322C^. (I) Cumulative plot of mIPSC amplitudes. The graphs (C), (E), (H) show values from individual neurons as dots and the mean values as bars with SEM as error bars. Statistical test applied was Kruskal-Wallis with post-hoc Dunn’s correction for multiple comparisons (**p≤0.01, ***p≤0.001, ****p≤0.0001).

**Figure 3:**
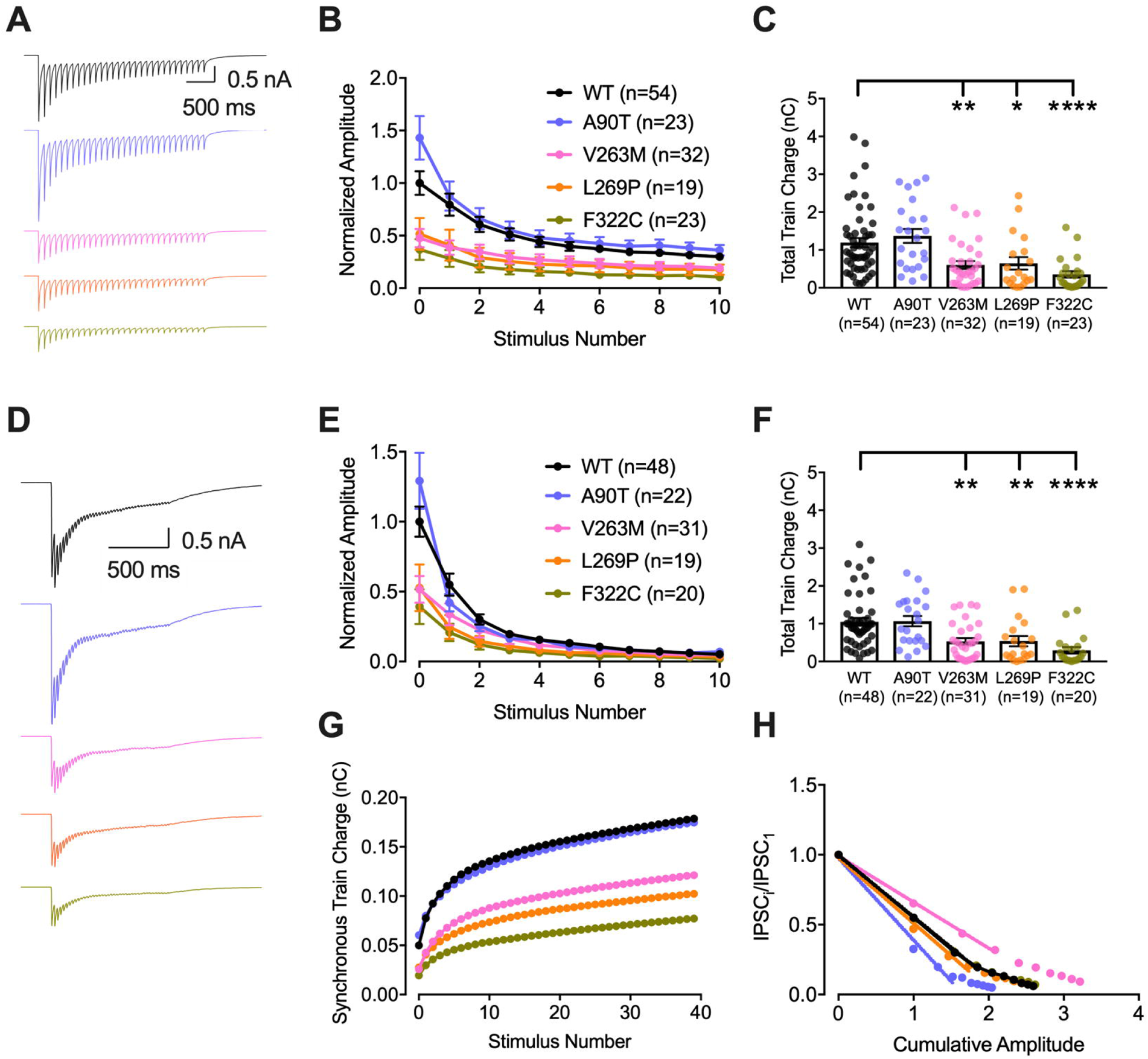
Stimulation with high frequency AP trains – VGAT^A90T^affects short term plasticity. (A) Averaged traces obtained during stimulation at 10 Hz from autaptic striatal *Slc32a1*^KO^ neurons with lentiviral expression of VGAT^WT^ (black), VGAT^A90T^ (blue), VGAT^V263M^ (pink), VGAT^L269P^ (orange), or VGAT^F322C^ (green). (B) Reduced IPSC amplitudes at 10Hz in neurons expressing VGAT^V263M^, VGAT^L269P^, VGAT^F322C^. Neurons expressing VGAT^A90T^ show stronger depression of the second amplitude. (C) Total charge of the GABAergic postsynaptic current during the 10 Hz train is reduced for VGAT^V263M^, VGAT^L269P^ and VGAT^F322C^. (D) Averaged traces obtained during stimulation at 40 Hz. (E) Reduced IPSC amplitudes at 40 Hz in neurons expressing VGAT^V263M^, VGAT^L269P^, VGAT^F322C^. Neurons expressing VGAT^A90T^ showed stronger depression of the second amplitude. (F) Total charge of the GABAergic postsynaptic current during the 40 Hz train is reduced for VGAT^V263M^, VGAT^L269P^ or VGAT^F322C^. (G) Cumulative synchronous IPSC charge of the 40Hz train. (H) Normalized Elmqvist-Quastel (EQ) plot of the first 10 responses to the 40Hz train. A line fit through the first four data points reveals that the release probability early in the train, *p*_EQ_, deviates from VGAT^WT^ for both VGAT^A90T^ and VGAT^V263M^, albeit in opposite directions. The graphs (C) and (F) show values from individual neurons as dots and the mean values as bars with SEM as error bars. Statistical test applied was Kruskal-Wallis with post-hoc Dunn’s correction for multiple comparisons (*p≤0.05, **p≤0.01,****p≤0.0001).

**Figure 4:**
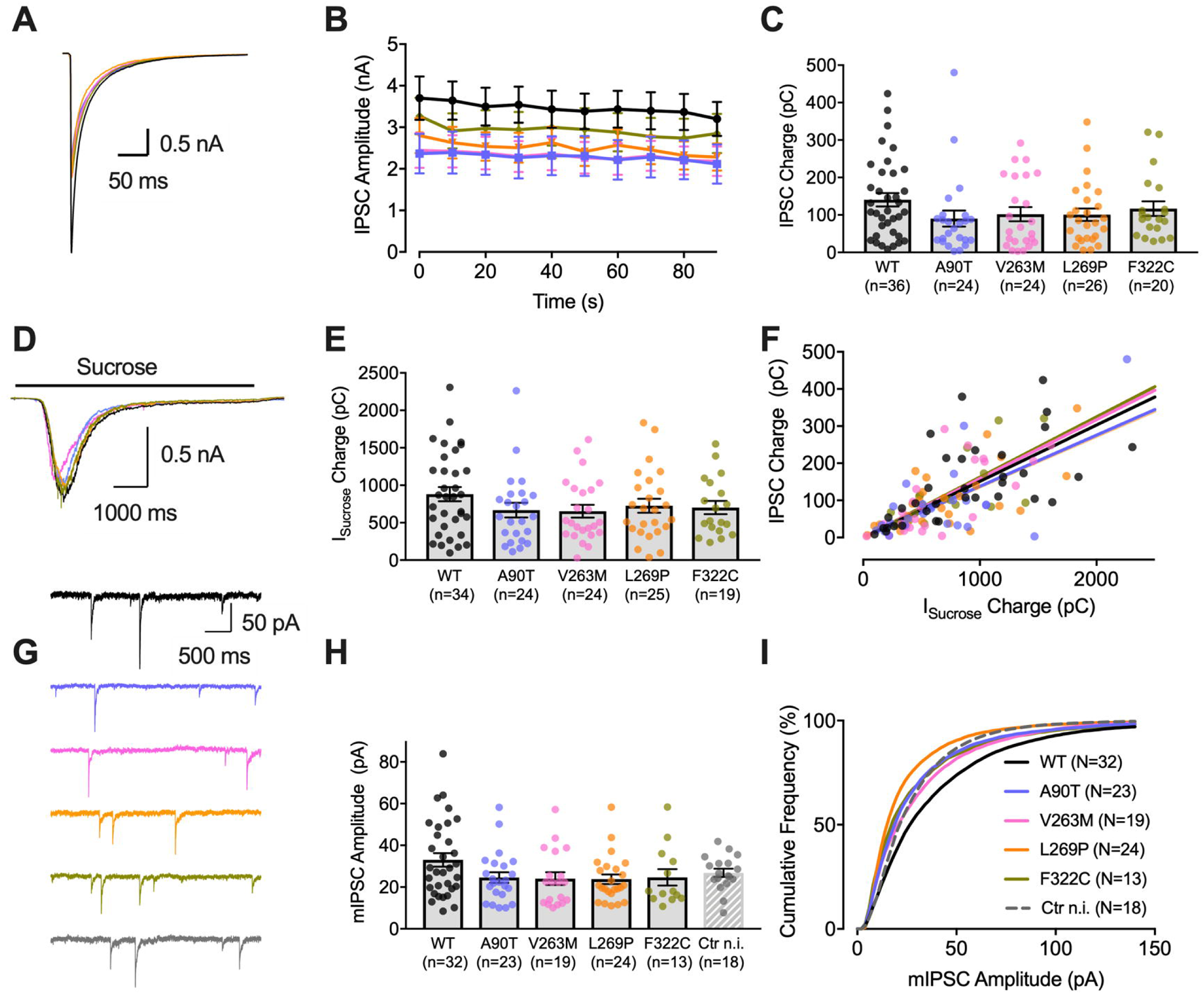
GABAergic IPSCs and mIPSCs in the presence of endogenous VGAT. (A) Example traces of initial, evoked inhibitory postsynaptic currents (IPSCs) from autaptic striatal *Slc32a1*^WT^ and *Slc32a1*^Het^ (*Slc32a1*^WT/Het^) neurons with lentiviral expression of VGAT^WT^ (black), VGAT^A90T^ (blue), VGAT^V263M^ (pink), VGAT^L269P^ (orange), or VGAT^F322C^ (green). (A) Average IPSC amplitudes at 0.1 Hz tend to be smaller for *Slc32a1*^WT/Het^ neurons expressing additional VGAT^A90T^, VGAT^V263M^, VGAT^L269P^, or VGAT^F322C^, but are not significantly reduced compared to neurons expressing VGAT^WT^. (C) IPSC charge is not significantly altered in *Slc32a1*^WT/Het^ neurons expressing any of the VGAT variants. (D) Averaged traces during the application of 0.5 M sucrose which triggers release of all fusion-competent SVs. (E) I_sucrose_ is not significantly changed by any of the variants in the presence of endogenous VGAT. (F) Average vesicular release probability 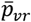, based on the linear fit of IPSC charge vs. I_sucrose_ charge, was not affected (G) Example traces of mIPSCs, representing the release of single SVs. Uninfected *Slc32a1*^WT^ and *Slc32a1*^Het^ neurons (Ctr n.i.) are shown in gray. (H) Average mIPSC amplitudes are not significantly reduced in neurons expressing VGAT^A90T^, VGAT^V263M^, VGAT^L269P^ or VGAT^F322C^ in addition to endogenous VGAT, although they tend to be smaller than mIPSC amplitudes in neurons expressing additional VGAT^WT^. (I) The cumulative mIPSC amplitude histogram shows a shift towards larger amplitudes for neurons expressing additional VGAT^WT^. The graphs (C), (E), (H) show values from individual neurons as dots and the mean values as bars with SEM as error bars. Statistical test applied was Kruskal-Wallis with post-hoc Dunn’s correction for multiple comparisons.

## Results

### Clinical description

Detailed clinical data on all four individuals are presented in Table 1 as well as in the Supplemental case reports and Supplemental Table 1. All four individuals showed global developmental delay and intellectual disability of varying degrees: moderate to severe in individuals 1, 2 and 4, while the respective information on individual 3 was not available. Early onset seizures were present in all four individuals, starting within the first 18 months of life with onset documented at (this information has been removed from the preprint version to comply with medrxiv policy). Seizure types consisted of complex partial seizures in individuals 1 and 4, myoclonic epilepsy with tonic-clonic seizures with status epilepticus in individual 2 and tonic-clonic seizures with fever in individual 3. Response to antiepileptic drugs was poor in individuals 1, 2 and 4. In contrast, individual 3 has been seizure free since the age of (this information has been removed from the preprint version to comply with medrxiv policy) with a therapy of valproate. Individuals 2, 3 and 4 showed choreatic, chorea-like, dystonic or dyskinetic movement phenotypes. Cranial imaging of individuals 3 and 4 showed normal results. The brain MRI of individual 1 revealed ventriculomegaly, loss of white matter and thinning of the corpus callosum. In individual 2, cerebral atrophy was noted.

**Table 1.**
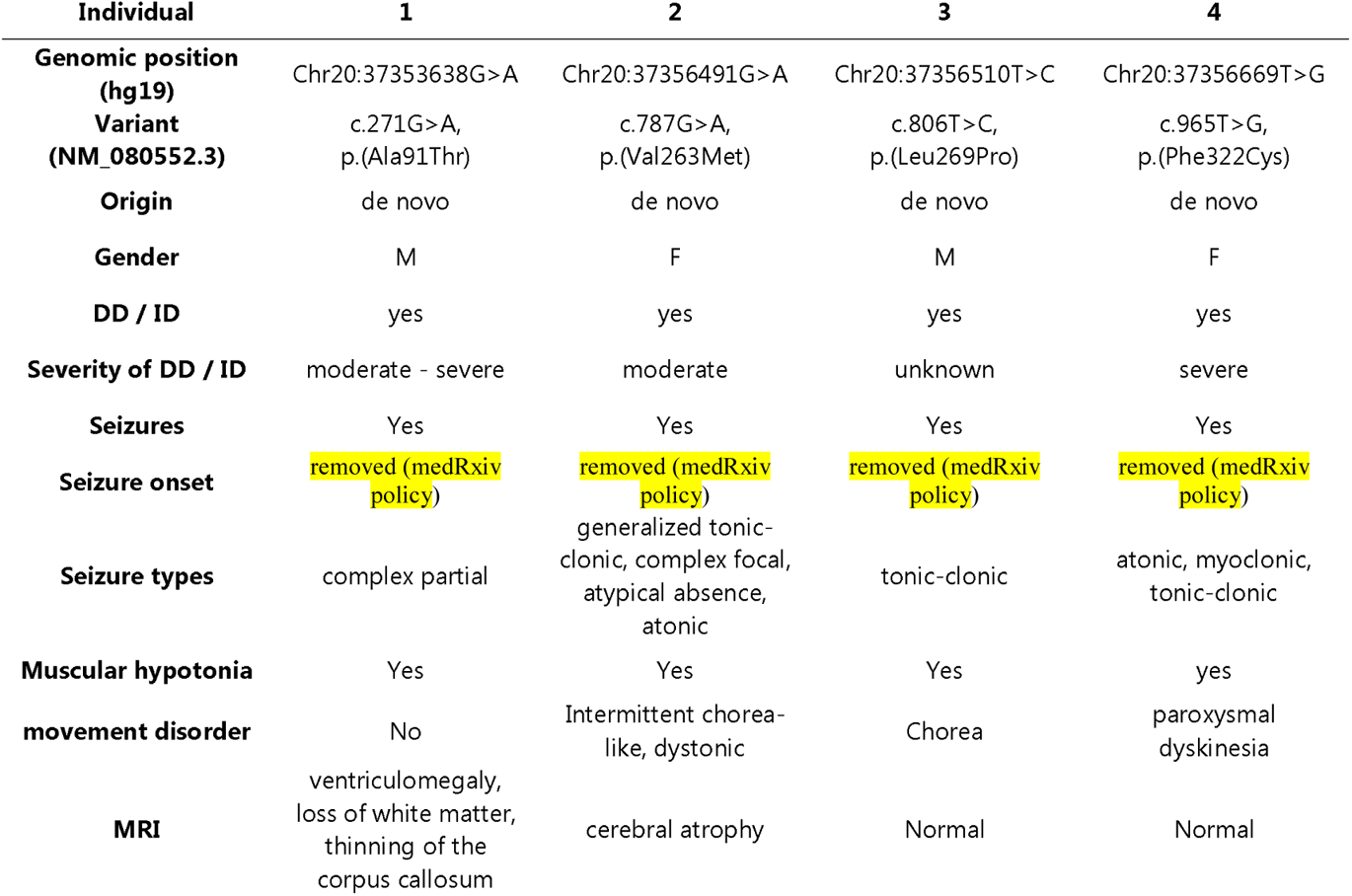
Summary of clinical symptoms of individuals with de novo variants in *SLC32A1*. Abbreviations: DD – developmental delay; ID – intellectual disability; F – female; M – male; Further clinical details are provided in Table S1.

| Individual | 1 | 2 | 3 | 4 |
| --- | --- | --- | --- | --- |
| **Genomic position (hg19)** | Chr20:37353638G>A | Chr20:37356491G>A | Chr20:37356510T>C | Chr20:37356669T>G |
| **Variant (NM_080552.3)** | c.271G>A,<br>p.(Ala91Thr) | c.787G>A,<br>p.(Val263Met) | c.806T>C,<br>p.(Leu269Pro) | c.965T>G,<br>p.(Phe322Cys) |
| **Origin** | de novo | de novo | de novo | de novo |
| **Gender** | M | F | M | F |
| **DD / ID** | yes | yes | yes | yes |
| **Severity of DD / ID** | moderate - severe | moderate | unknown | severe |
| **Seizures** | Yes | Yes | Yes | Yes |
| **Seizure onset** | removed (medRxiv policy) | removed (medRxiv policy) | removed (medRxiv policy) | removed (medRxiv policy) |
| **Seizure types** | complex partial | generalized tonic-clonic, complex focal, atypical absence, atonic | tonic-clonic | atonic, myoclonic, tonic-clonic |
| **Muscular hypotonia** | Yes | Yes | Yes | yes |
| **movement disorder** | No | Intermittent chorea-like, dystonic | Chorea | paroxysmal dyskinesia |
| **MRI** | ventriculomegaly, loss of white matter, thinning of the corpus callosum | cerebral atrophy | Normal | Normal |

### Causative variants

All four *de novo* variants in *SLC32A1* identified in this study comprise missense changes (Fig. 1). All variants are absent from gnomAD. Variants in individuals 2, 3 and 4 affect highly conserved amino acids and are predicted to be damaging by multiple *in silico* prediction tools (Table S2). All three variants are located in helices that form the substrate transport pathway. The *de novo* variant in individual 1 affects a weakly conserved amino acid and is predicted to be benign by most *in silico* prediction tools.

**Figure 1:**
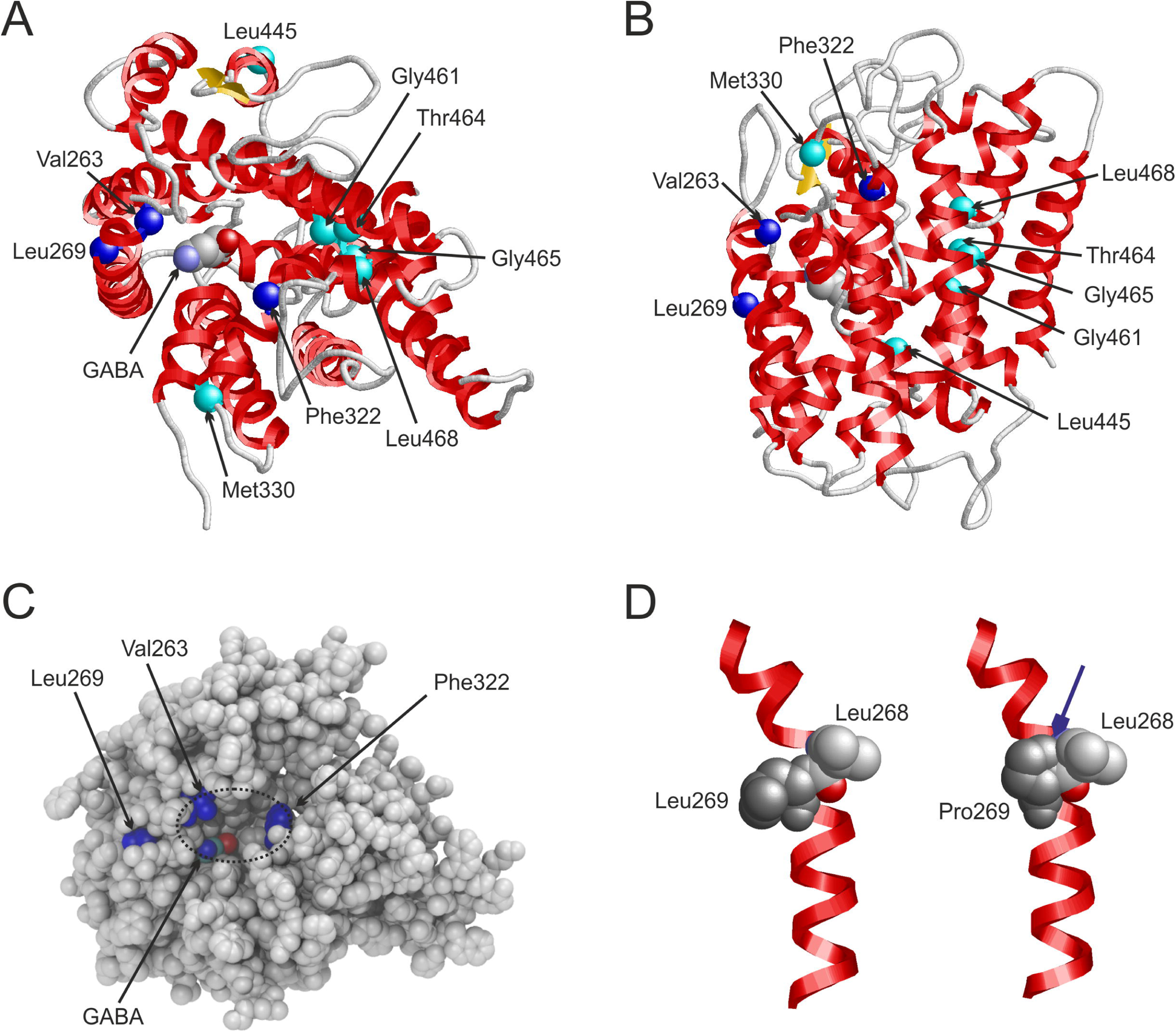
Model of the VGAT transmembrane domain (A) View on VGAT from the cytosolic side indicating the sites of sequence variants. Positions described in the present study are highlighted as blue spheres, whereas additional positions identified in the previous study by Heron et al.^5^ are marked as cyan spheres. The structure is shown in ribbon presentation with helices highlighted in red and the substrate GABA in space-filled presentation (atom-type coloring). (B) Side view on VGAT, which is rotated by 90° around the horizontal axis compared to the view in (A). (C) View from the cytosolic side on VGAT in space-filled presentation (same orientation as in (A)). The dotted circle marks a cavity that is lined by Val263 and Phe322. The position of a GABA molecule at the bottom of the cavity is also indicated. (D) Leu269 is located in one of the transmembrane helices (left). A proline at this position (right) will cause steric clashes (marked by an arrow) with the adjacent Leu268 thereby affecting the helix conformation and stability.

### Structural modelling

A VGAT model, comprising the transmembrane domain (residues 115-509) of the transporter, allowed us to investigate the structural consequences of the variants p.(Val263Met), p.(Leu269Pro) and p.(Phe322Cys) in more detail. All three missense variants are located in helices that line the substrate transport pathway (Fig. 1AB).

A closer view of the VGAT structure, which represents the cytosol-open state of the transporter, revealed that residues Val263 and Phe322 line a deep cavity with the GABA substrate at its bottom (Fig. 1C). The variants p.(Val263Met) and p.(Phe322Cys) are predicted to cause a significant (≥70 Å^3^) change in the size of this cavity, indicating that they likely affect substrate uptake on the cytosolic side or the transport process itself. The fact that Val263 and Phe322 are accessible from the cytosolic side (Fig. 1C) indicates that mutating these sites might additionally affect binding of cytosolic interaction partners to VGAT. Leu269 is located in an α−helix that flanks the central cavity. An exchange to proline causes steric interference with the adjacent Leu268 (Fig. 1D), which destabilizes the structure close to the site of substrate transport.

### Functional analyses

To examine the effects of the four VGAT variants on inhibitory neurotransmission, we expressed lentiviral constructs of each variant in striatal GABAergic mouse neurons in microisland cultures. Single neurons growing on astrocyte microislands only form synapses with themselves. Electrophysiological recordings from such isolated, autaptic neurons allow for the detailed analysis of neurotransmission of individual neurons under defined conditions and are ideally suited for structure-function analyses of synaptic proteins.^2, 25, 31–33^

Human and murine VGAT show 98.5% identity at the amino acid level. The majority of amino acid exchanges between species can be found in the N-terminal region, where the p.(Ala91Thr) variant is located. Because we hypothesized that this variant may disrupt protein-protein interactions, and/or trafficking of VGAT^34^, we chose to introduce this and all other variants into lentiviral expression constructs of the murine VGAT cDNA, rather than work with human VGAT in mouse neurons. The p.(Ala91Thr) variant corresponds to VGAT^A90T^ in mouse, whereas the amino acid positions of all other variants are conserved. We produced lentiviral particles which express VGAT and EGFP from separate promoters. Expression of full-length VGAT protein was confirmed by Western blotting of infected striatal *Slc32a1*^KO^ neurons in mass culture (Fig. S1). We first evaluated the effects on GABAergic neurotransmission in the absence of endogenous wild-type (WT) VGAT protein, by analyzing to what degree each variant was able to restore GABAergic neurotransmission to mouse VGAT knockout (*Slc32a1*^KO^) neurons. Without rescue by exogenous VGAT expression, the majority of inhibitory *Slc32a1*^KO^ neurons are synaptically completely silent and the remainder show only residual neurotransmission.^2^ For electrophysiological recordings, we infected autaptic striatal cultures of *Slc32a1*^KO^ neurons on DIV 2-3. Infected neurons were identified based on EGFP expression and phenotypic rescue of GABAergic neurotransmission in *Slc32a1*^KO^ neurons was assessed between DIV 9-14 by patch-clamp recordings in whole-cell voltage clamp configuration.

### Reduced evoked GABAergic neurotransmission in neurons expressing VGAT^V263M^, VGAT^L269P^, or VGAT^F322C^

We first assayed basal synaptic strength of GABAergic autapses by recording single action potential (AP)-evoked inhibitory postsynaptic currents (IPSCs) at an inter-stimulus interval of 10 s. IPSCs were elicited by 1 ms step depolarizations, which produce single escaping APs and trigger Ca^2+^-dependent SV exocytosis. *Slc32a1*^KO^ neurons expressing VGAT^V263M^, VGAT^L269P^, or VGAT^F322C^, generated strongly reduced IPSCs when compared to *Slc32a1*^KO^ neurons rescued with wild-type VGAT (VGAT^WT^) (Fig. 2A-C). IPSC amplitude (Fig. 2B) and IPSC charge (Fig. 2C) were significantly smaller for these three variants. IPSCs in neurons expressing VGAT^A90T^ on the other hand, tended to be slightly larger than VGAT^WT^ IPSCs, although the difference was statistically not significant (Fig. 2A-C, Table 2). Total cell capacitance, which is a measure of cell size and complexity of dendritic arborization, was not affected by expression of any of the variants (Table 2), indicating that growth and differentiation of neurons was unaffected.

Single AP-induced IPSCs typically consume only a small fraction of fusion-competent SVs. To estimate the maximum inhibitory synaptic strength, i.e. the product of the total number of readily releasable SVs and quantal size, GABA release was elicited by bath application of 0.5 M sucrose solution for 7 s (Fig. 2D,E). Such hypertonic stimuli generate transient postsynaptic current responses (I_sucrose_) reflecting exocytosis of all fusion-competent SVs in a Ca^2+^-independent manner. I_sucrose_ therefore represents an estimate of the maximum synaptic inhibition achievable during high-frequency AP bursts, which lead to rapid consumption of nearly all fusion-competent SVs. Compared to *Slc32a1*^KO^ neurons expressing VGAT^WT^, I_sucrose_ was significantly reduced in neurons expressing VGAT^V263M^, VGAT^L269P^, or VGAT^F322C,^ but not in neurons expressing VGAT^A90T^ (Fig. 2E, Table 2). We then estimated the average vesicular release probability 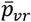 representing the fraction of fusion-competent SVs released during single APs, by fitting regression lines to scatter plots of IPSC charge vs. I_sucrose_ charge (Fig. 2F). Surprisingly, 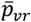 was significantly elevated in *Slc32a1*^KO^ neurons expressing VGAT^A90T^ but very similar to VGAT^WT^ in neurons expressing VGAT^V263M^, VGAT^L269P^, or VGAT^F322C^ (Fig. 2F, Table 2).

### Reduced quantal size in neurons expressing VGAT^V263M^, VGAT^L269P^ or VGAT^F322C^

To assess whether the reduction in evoked GABAergic neurotransmission can be attributed to reduced vesicular GABA content, we next evaluated the quantal size by measuring amplitudes of miniature inhibitory postsynaptic currents (mIPSCs), which correspond to the spontaneous fusion of single SVs (Fig. 2G-I). Because mIPSC frequency was generally low in resting neurons of striatal autaptic cultures, we recorded mIPSCs during the recovery periods following brief 10 Hz stimulation (30 APs, see below). During repetitive AP firing, presynaptic intracellular Ca^2+^ transiently rises, thereby increasing the rate of asynchronous or delayed SV fusion events. The mIPSC amplitude distribution in *Slc32a1*^KO^ neurons expressing VGAT^A90T^ was indistinguishable from that of neurons expressing VGAT^WT^ (Fig 2I). By contrast, cumulative probability plots of mIPSC amplitudes showed a marked left shift for VGAT^V263M^, VGAT^L269P^, and VGAT^F322C^ expressing neurons, indicating a general reduction in mIPSC size (Fig 2I). Accordingly, average mIPSC amplitudes were significantly reduced for these three variants, (Fig. 2H,I, Table 2 and Fig. S2), which is consistent with less GABA being loaded into SVs carrying these transporter variants. *Slc32a1*^KO^ neurons expressing VGAT^V263M^, VGAT^L269P^, or VGAT^F322C^ not only exhibited reduced mIPSC size, but their average mIPSC frequencies also tended to be lower (Table 2). Because partially filled and even empty SVs are known to fuse,^31^ an unknown fraction of very small mIPSCs, representing fusion of partially filled SVs, may escape detection. Therefore, the average quantal size measured in VGAT^V263M^, VGAT^L269P^, and VGAT^F322C^ expressing neurons is likely an overestimate. Furthermore, undetected SV fusion events are likely to at least partially account for the reduced mIPSC frequencies observed in neurons expressing VGAT^V263M^, VGAT^L269P^, or VGAT^F322C^ (Table 2).

Immediately after stimulus trains, mIPSC frequencies were strongly elevated, sometimes leading to partial overlap and temporal summation of individual events, which not only impairs automatic mIPSC detection via template matching, but also complicates the analysis of mIPSC kinetics. We therefore additionally recorded mIPSCs under conditions of slightly elevated bath osmolarity (50 mM sucrose), which increased the average rate of spontaneously occurring mIPSCs relative to resting conditions but resulted in fewer overlapping events compared to mIPSC recordings immediately after AP firing. Analysis of the kinetic properties of mIPSCs recorded under conditions of elevated bath osmolarity revealed significantly slower rise times for VGAT^V263M^, VGAT^L269P^, and VGAT^F322C^ (Table 2), consistent with a lower peak concentration of GABA being released into the synaptic cleft. Decay times were not significantly different from VGAT^WT^, although they tended to be slower for VGAT^V263M^, VGAT^L269P^, and VGAT^F322C^, which may be due to the less favorable signal to noise ratio of the smaller mIPSCs recorded in these cells as compared to *Slc32a1*^KO^ neurons expressing VGAT^WT^ or VGAT^A90T^. In summary, GABAergic neurons expressing VGAT^V263M^, VGAT^L269P^, or VGAT^F322C^ show reduced mIPSC quantal size, which is consistent with lower vesicular GABA content, whereas this was not the case for VGAT^A90T^.

### Altered short-term plasticity in neurons expressing VGAT^A90T^ or VGAT^V263M^

Thus far, we had assayed synaptic strength in response to single presynaptic APs. Because inhibitory synapses in intact circuits are typically activated by AP bursts, we recorded synaptic responses elicited by short AP trains to evaluate how well *Slc32a1*^KO^ neurons expressing VGAT^A90T^, VGAT^V263M^, VGAT^L269P^, or VGAT^F322C^ can sustain inhibitory neurotransmission at higher AP firing rates in *Slc32a1*^KO^ neurons (Fig. 3). Repetitive stimulation at 10 Hz (Fig. 3A-C) and 40 Hz (Fig. 3D-H) leads to a gradual decline of IPSC amplitudes (Fig. 3B, Fig. 3E). In addition to the synchronous IPSCs, postsynaptic responses also consisted of an asynchronous release component, most likely triggered by the sustained elevation in intracellular presynaptic [Ca^2+^].^35, 36^ The total IPSC train charge (IPSC integral over 4.25 s for 10 Hz and over 2.25 s for 40 Hz) representing the sum of synchronous and asynchronous GABAergic neurotransmission was significantly reduced for VGAT^V263M^, VGAT^L269P^, and VGAT^F322C^ but similar to VGAT^WT^ for neurons expressing VGAT^A90T^ (Fig. 3C, and Fig. 3F). To test whether synchronous and asynchronous release are similarly affected by the VGAT variants, we obtained an estimate for the number of synchronously released quanta (m_i_) by each train stimulus i by the amplitude ratio IPSC_i_/mIPSC and plotted the synchronous IPSC train charge (m_i_ × q) in Fig. 3G. For a comparison of synchronous IPSC waveforms with that of total IPSCs, we convolved trains of impulse responses scaled by m_i_ with the respective mIPSC waveforms (Fig. S3). Both types of analysis (Fig. 3G, Fig. S3) indicate that the relative contribution of synchronous and asynchronous GABA release to the total IPSCs was unaffected by any of the VGAT variants.

Remarkably, *Slc32a1*^KO^ neurons expressing VGAT^A90T^ showed an altered short-term plasticity in response to 10 Hz and 40 Hz stimulus trains. Consistent with their higher 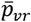 (Fig. 2F; Table 2), neurons expressing VGAT^A90T^ exhibited larger initial IPSCs and more rapid synaptic depression in response to 10 Hz and 40 Hz stimulus trains (Fig. 3B, Fig. 3E).

Elevated 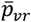 is expected to cause faster consumption of fusion-competent SVs during AP trains resulting in more rapid synaptic depression, whereas neurons with low 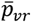 may show facilitation, i.e. a transient enhancement of amplitudes. Among cultured VGAT^WT^-expressing *Slc32a1*^KO^ neurons, cells showing either only IPSC depression or transient IPSC facilitation were both observed. By contrast, *Slc32a1*^KO^ neurons expressing VGAT^A90T^ rarely showed IPSC facilitation. Average IPSC waveforms for all cells analyzed are shown in Fig. 3A and Fig. 3D. To corroborate elevated 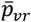 in neurons expressing VGAT^A90T^, we plotted the amplitude decrement during the first four IPSCs (IPSC_n_) measured during 40 Hz trains versus the cumulative amplitude of all previous IPSCs (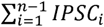) (Fig. 3H). Under conditions of negligible SV pool replenishment during inter-stimulus intervals, the negative slope of such plots represents a measure of release probability (*p*_EQ_).^38^ In contrast to the 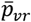, estimated by the linear fit of IPSC charge vs I_sucrose_ charge (Fig. 2F), *p*_EQ_ is dominated by SVs fusing early during IPSC trains and may therefore be slightly higher if release probability is not homogeneous among all fusion-competent SVs.^39^ *P*_EQ_ was higher for VGAT^A90T^ expressing cells than for cells expressing VGAT^WT^ or any of the other variants (Fig. 3H, Table 2). Surprisingly, the slope of the line fit also deviated for *Slc32a1*^KO^ neurons expressing VGAT^V263M^ compared to those expressing VGAT^WT^, albeit in the opposite direction, indicating a lower *p*_EQ_ for *Slc32a1*^KO^ neurons expressing VGAT^V263M^ (Fig. 3H, Table 2), even though their 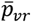 was unaltered when compared to neurons expressing VGAT^WT^ (Fig. 2F, Table 2).

### No dominant-negative effects on GABAergic neurotransmission in the presence of endogenous VGAT

Thus far, we had investigated the intrinsic properties of each VGAT variant in *Slc32a1*^KO^ neurons, i.e. in the absence of endogenous VGAT. However, since all patients are heterozygous for their respective variant, we wanted to assess if any of the variants might exert a dominant-negative effect in the presence of a *Slc32a1*^WT^ allele. Since co-infection with multiple viruses is inefficient, we decided to infect neuronal cultures from embryos either heterozygous (*Slc32a1*^Het^), or homozygous for the endogenous VGAT allele (*Slc32a1*^WT^). Since no obvious differences were detected between these two genotypes upon lentiviral expression of the VGAT variants, we pooled the data obtained in WT and Het cultures (*Slc32a1*^WT/Het^). In the presence of endogenous VGAT, additional lentiviral expression of VGAT^WT^, or any of the variants (VGAT^A90T^, VGAT^V263M^, VGAT^L269P^, VGAT^F322C^) did not cause any differences in basic GABAergic neurotransmission that were statistically significant (Fig. 4 and Table 2). Although we observed a tendency towards smaller IPSC amplitudes (Fig. 4A,B) and IPSC charge (Fig. 4C) for all four VGAT variants compared to *Slc32a1*^WT/Het^ neurons infected with VGAT^WT^-encoding lentivirus, we suspected that this may be due to the fact that overexpression of vesicular transporters can increase quantal size beyond what is observed in uninfected control cells.^31, 40, 41^ We therefore included uninfected *Slc32a1*^WT/Het^ control neurons (Ctr n.i.) in our analysis of mIPSC amplitudes (Fig. 4G-I). Average quantal size was not significantly different for any of the variants and similar to uninfected control neurons, whereas neurons expressing VGAT^WT^ tended to have slightly larger mIPSC amplitudes (Fig. 4H). The cumulative mIPSC amplitude distribution (Fig. 4I) and mIPSC amplitude histogram (Fig. S4) of VGAT^WT^ expressing neurons were shifted towards larger amplitudes in comparison to all four VGAT variants, and also compared to uninfected control neurons. Taken together, these findings indicate that VGAT^A90T^,VGAT^V263M^, VGAT^L269P^ and VGAT^F322C^ are less effective at boosting quantal size than VGAT^WT^ when expressed in the presence of the *Slc32a1*^WT^ allele, but do not exert a dominant-negative effect.

An unexpected result in this set of experiments is that in the presence of endogenous VGAT, VGAT^A90T^ appeared to perform more similarly to the other variants with respect to IPSC amplitude, IPSC charge, I_sucrose_ charge and quantal size. Since VGAT^A90T^ was able to rescue quantal size as effectively as VGAT^WT^ when expressed in KO neurons (Fig. 2H,I), we would have expected VGAT^A90T^ to be able to boost quantal size in the presence of endogenous VGAT. This, however, was not the case (Fig. 4H,I), which indicates, that VGAT^A90T^ may perform slightly differently in the presence of VGAT^WT^ than in its absence.

## Discussion

In this study we describe in detail four individuals with a novel neurodevelopmental disorder with epilepsy that harbor *de novo* missense variants in *SLC32A1*. Reduced GABAergic neurotransmission due to impaired SV loading is the likely cause of the developmental and the epilepsy phenotype in three of the affected individuals, and we provide evidence that effects on synaptic short-term plasticity may be involved in the fourth individual.

According to data from gnomAD, *SLC32A1* is a gene with a reduced number of missense variants in controls, indicating a selective constraint on this type of variant in a control population that is not affected by severe, early-onset phenotypes such as epilepsy, DD and ID (z score for missense variants = 2.3, observed/expected ratio = 0.63).^14^ *De novo* occurrence, strong *in silico* and functional data support the causality of variants in individuals 2, 3 and 4 that are located in the transporter domain and cause reduced GABAergic neurotransmission, consistent with impaired SV filling. While the evidence for causality in individual 1 is less compelling, the functional deficits in synaptic short-term plasticity identified in cultured *Slc32a1*^KO^ neurons expressing the N-terminal variant VGAT^A90T^ were quite striking, and constitute a likely pathomechanism. Since GABAergic neurons often signal via high frequency AP bursts,^42^ stronger synaptic depression may affect signal processing at individual connections as well as in neuronal circuits. Furthermore, we also obtained evidence for alterations in release probability in neurons expressing VGAT^V263M^ when estimated from early synaptic depression during 40 Hz trains. Distinct effects on *p* and therefore synaptic short-term plasticity have previously been demonstrated for the vesicular glutamate transporters VGLUT1 and VGLUT2. In the case of VGLUT1, lower *p* was shown to be due to an interaction between VGLUT1 and endophilin A1, a regulator of endocytosis.^43^ The VGAT N-terminus contains several motifs that could be involved in endocytic sorting and activity dependent recycling, and Alanine 91 (Alanine 90 in mouse) lies adjacent to a polyproline motif of unknown function.^34^ However, since VGAT^A90T^ effectively rescued quantal size in *Slc32a1*^KO^ neurons, this variant appears to be correctly sorted to SVs, at least in the absence of an *Slc32a1*^WT^ allele. Nonetheless, variants in the N-terminal domain could potentially affect *p* by interfering with VGAT or SV recycling or with other SV properties. Interestingly, Heron et al.^5^ also describe an N-terminal VGAT variant, p.Gly43Cys, which is in fact located within an SV targeting motif identified by Santos et al.^34^

Within the VGAT transport domain, the location of the variants predicted to affect substrate transport that we identify in this study, are clearly distinct from most variants reported previously by Heron et al^5^ in affected individuals with GEFS+ and familial epilepsy, which are located more distal to the substrate transport domain. This difference may explain why the latter group of variants resulted in a milder phenotypic presentation of the affected individuals, especially without DD/ID. A pronounced deficit in VGAT transporter function could potentially even influence trophic actions of GABA early in development, or the developmental shift of GABAergic neurotransmission from excitation to inhibition during nervous system maturation.^44–46^ The amino acid Valine at position 263 is the only position where multiple variants were found in affected individuals. Interestingly, the aforementioned alteration of the cavity size with the GABA substrate at its bottom is smaller for the previously reported variant p.(Val263Ala)^5^ compared to the variant p.(Val263Met) identified in this study. This, as well as the apparent effect on *p* seen in our study, could potentially explain the distinct and more severe phenotypic effects observed for this variant, while the affected individuals identified by Heron et al.^5^ did not show DD/ID.

We did not find evidence for altered pain tolerance in the affected individuals in our cohort – a symptom that might point toward disrupted glycinergic or also GABAergic transmission due to the impaired transport capacity of VGAT in neurons of the spinal cord. There is very little expression of VGAT outside the nervous system, and we did not identify any prominent non-neurological symptoms in the affected individuals in our cohort.^47^

GABAergic neurotransmission has long been known to play a causal role in epileptic phenotypes as variants in a variety of GABA receptor subunits were shown to be causal, especially *de novo* variants in early onset developmental and epileptic encephalopathy (DEE).^4, 48^ The phenotype of the patients in the present study is in line with what is known about disrupted GABA receptor function caused by *de novo* variants. The wide phenotypic variability ranging from milder epilepsy syndromes such as a GEFS+ phenotype and familial epilepsies, to the severe end of the spectrum with the phenotype of DEE, has also been shown to be caused by disrupted GABA receptor function, e.g. due to variants in *GABRA1*, *GABRG2* or *GABRB3*.^48–50^

In summary, our findings establish *de novo* missense variants in *SLC32A1* as a cause of neurodevelopmental disorder that adds to the description of neuro-genetic disorders associated with impaired GABAergic transmission.

## Data Availability

All data produced in the present study are available upon reasonable request to the authors.

## Abbreviations

AP: action potential
DD: developmental delay
DIV: day in vitro
GEFS+: genetic epilepsy with febrile seizures plus
ID: intellectual disability
IGE: idiopathic generalized epilepsy
IPSC: inhibitory postsynaptic current
KO: knockout
mIPSC: miniature Inhibitory Postsynaptic Current
VGAT: vesicular GABA transporter
VIAAT: vesicular inhibitory amino acid transporter
WT: wild type

## Acknowledgements

We thank the families for their participation and support of this study. We would also like to thank JeongSeop Rhee and ChungKu Lee for generously providing astrocyte cultures, and Astrid Zeuch for excellent technical assistance.

## Web Resources

GnomAD, http://gnomad.broadinstitute.org/

GeneMatcher, https://genematcher.org/

GenBank, https://www.ncbi.nlm.nih.gov/genbank/

OMIM, http://www.omim.org/

