## Supplemental Data for "De novo missense variants in *SLC32A1* cause a neurodevelopmental disorder with epilepsy due to impaired GABAergic neurotransmission"

### Inhalt

### Supplemental case reports

**Individual 1: c.271G>A, p.(Ala91Thr)**

This case report has been removed from the preprint version to comply with medrxiv policy. Please see the published version or contact the authors if you are interested in this information.

Trio exome sequening in this individual identified a second de novo variant: Chr16(GRCh37):g.1252243, NM_021098.2:c.1793C>T, p.(Ala598Val) in the *CACNA1H* gene. The status of CACNA1H as a cause of monogenic cause of epilepsy has been controversial and a recent review of available data postulated there is only limited evidence for a causal role.[1]

**Individual 2:, c.787G>A, p.(Val263Met)**

This case report has been removed from the preprint version to comply with medrxiv policy. Please see the published version or contact the authors if you are interested in this information.

**Individual 3: c.806T>C, p.(Leu269Pro)**

This case report has been removed from the preprint version to comply with medrxiv policy. Please see the published version or contact the authors if you are interested in this information.

**Individual 4: c.965T>G, p.(Phe322Cys)**

This case report has been removed from the preprint version to comply with medrxiv policy. Please see the published version or contact the authors if you are interested in this information.

### Supplemental Tables

**Table S1.** Detailed clinical information of individuals with de novo variants in *SLC32A1* (Excel-file)

#### Table S2 In silico prediction of all de novo variants in *SLC32A1*.

| Indi-vidual | chr20: g.(hg19) | c. | p. | CADD 1.6 (Kircher et al.)[2] | REVEL (Ioannidis et al.)[3] | Mutation Taster2 (Schwarz et al.)[4] | M-CAP 1.3 (Jagadeesh et al.)[5] | Polyphen-2 v2.2.2 (Adzhubei et al.)[6] | GERP++ (Cooper et al.)[7] | Conservation | Absent from GnomAD (v2.1.1) |
|---|---|---|---|---|---|---|---|---|---|---|---|
| 1 | 37353638 | c.271G>A | p.(Ala91Thr) | 20.8 | 0.115 | DC | T | B | 4.97 | low, c. familiaris | yes |
| 2 | 37356491 | c.787G>A | p.(Val263Met) | 27.4 | 0.462 | DC | T | PrD | 4.87 | high, c. elegans | yes |
| 3 | 37356510 | c.806T>C | p.(Leu269Pro) | 29.9 | 0.414 | DC | D | PrD | 4.87 | high, x. laevis | yes |
| 4 | 37356669 | c.965T>G | p.(Phe322Cys) | 25.4 | 0.591 | DC | D | PrD | 4.73 | high, c. elegans | yes |

### Supplemental Figures

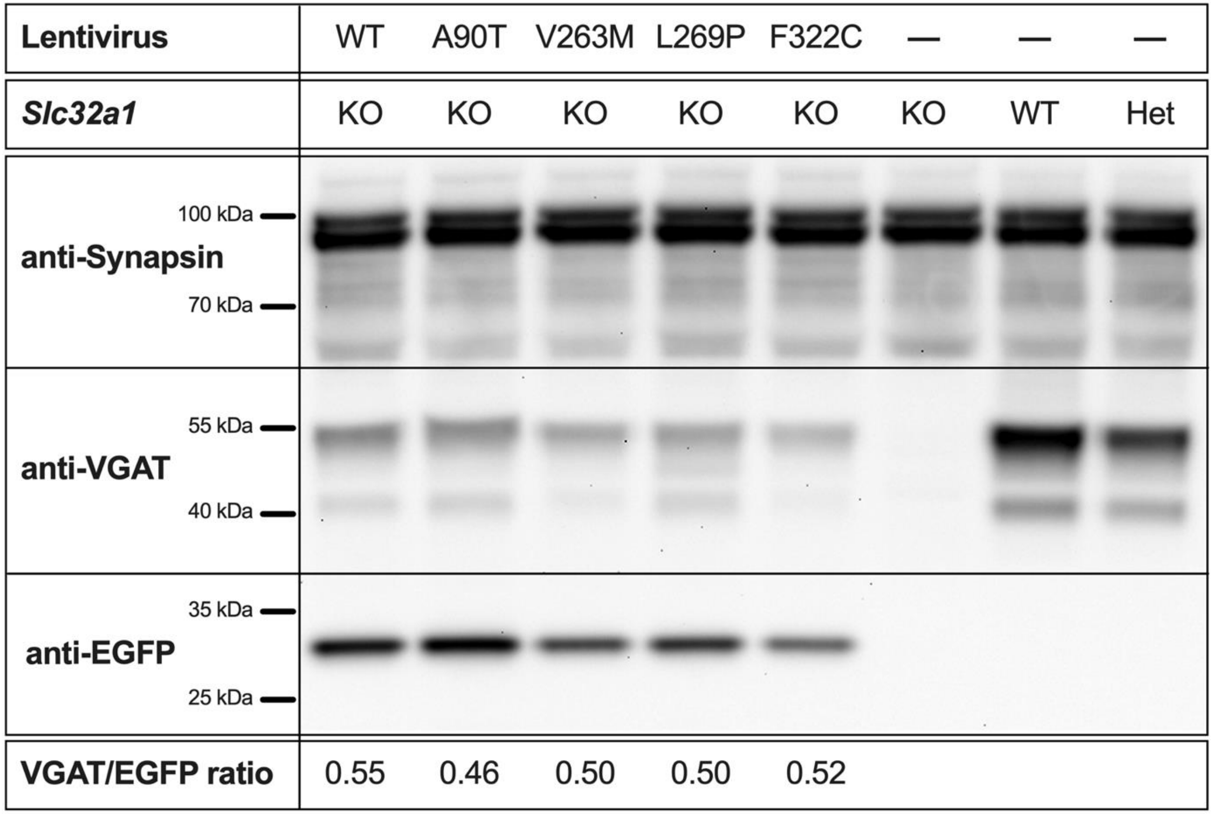

Platzer et al. Figure S1

Figure S1: Western blot of *Slc32a1*$^{KO}$ striatal mass cultures infected with lentiviral particles expressing the respective VGAT variants and EGFP from separate promoters. Uninfected *Slc32a1*$^{KO}$, *Slc32a1*$^{WT}$ and *Slc32a1*$^{Het}$ cultures were included as control (last 3 lanes). The VGAT signal of infected *Slc32a1*$^{KO}$ cultures (lanes 1-5) is weaker than the endogenous VGAT signal from *Slc32a1*$^{WT}$ and *Slc32a1*$^{Het}$ cultures (last 2 lanes), because lentiviral titers were not high enough to infect all neurons of the *Slc32a1*$^{KO}$mass culture. However, all lentiviral constructs produced full-length VGAT protein and VGAT/EGFP ratios were similar for all 5 lentiviral constructs, indicating similar expression of VGAT relative to the internal EGFP control.

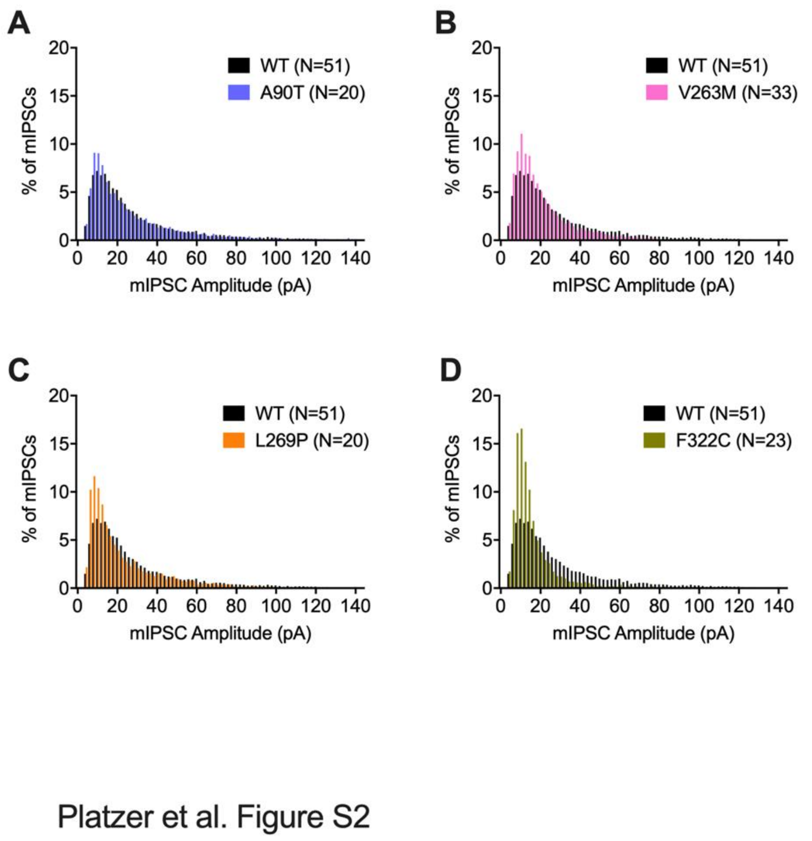

**Figure S2:** Comparison of mIPSC amplitude distributions for *Slc32a1*$^{KO}$ neurons rescued with (A) VGAT$^{A90T}$ vs VGAT$^{WT}$ (B) VGAT$^{V263M}$ vs VGAT$^{WT}$ (C) VGAT$^{L269P}$ vs VGAT$^{WT}$ (D) VGAT$^{F322C}$ vs VGAT$^{WT}$ .

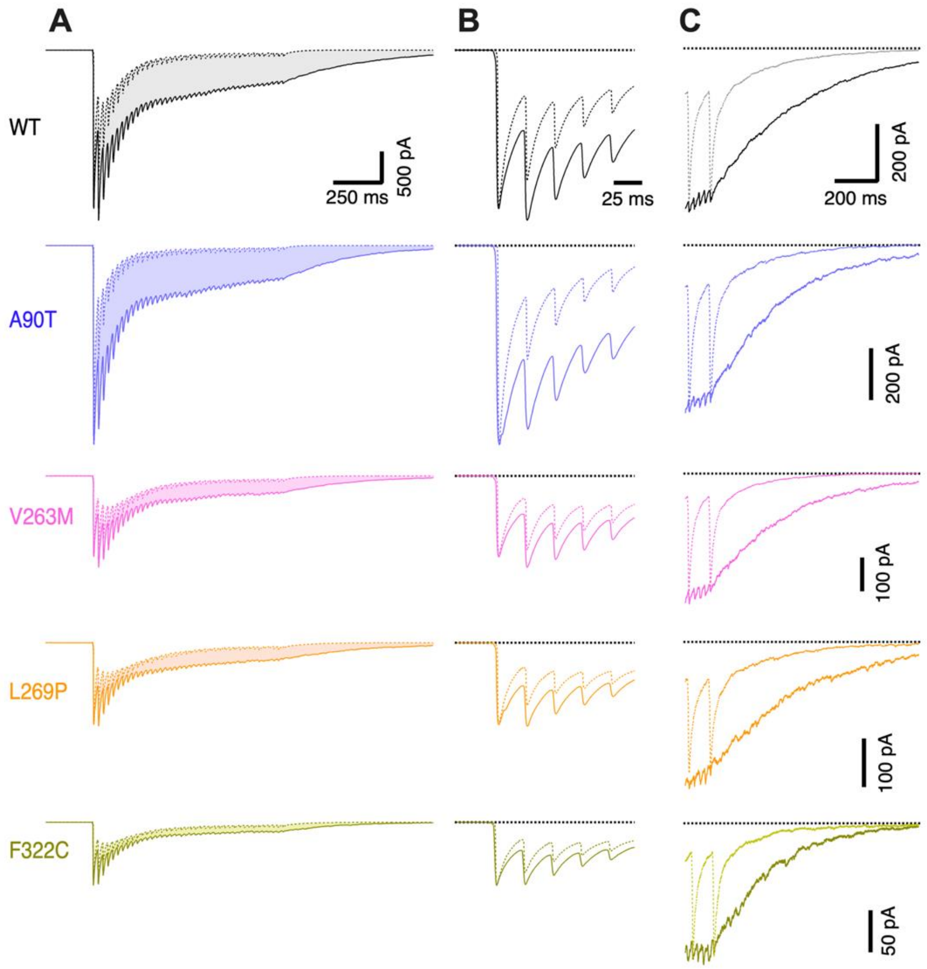

Platzer et al. Figure S3

**Figure S3:** Comparison of synchronous and total 40 Hz IPSC waveforms. Total IPSCs (solid traces) represent averages of peak-aligned IPSCs from all cells analyzed. (A) Estimates for the synchronous IPSC components (dotted traces) were obtained by convolving a train of impulse responses (scaled by the mean quantal content for each IPSC in the train) with the respective mIPSC waveform for each genotype. The shaded area represents an estimate for the asynchronous GABA release. (B) Initial five IPSCs of the average traces in (A) shown at a faster time scale. The relative contributions of synchronous and asynchronous release to the total 40 Hz IPSCs were comparable between genotypes. (C) Comparison of the decay of the last IPSC of the 40 Hz IPSC trains with a peak-scaled version of the last IPSC of the 10 Hz IPSC trains illustrating profoundly slowed IPSC decay following high-frequency stimulation due to delayed asynchronous release.

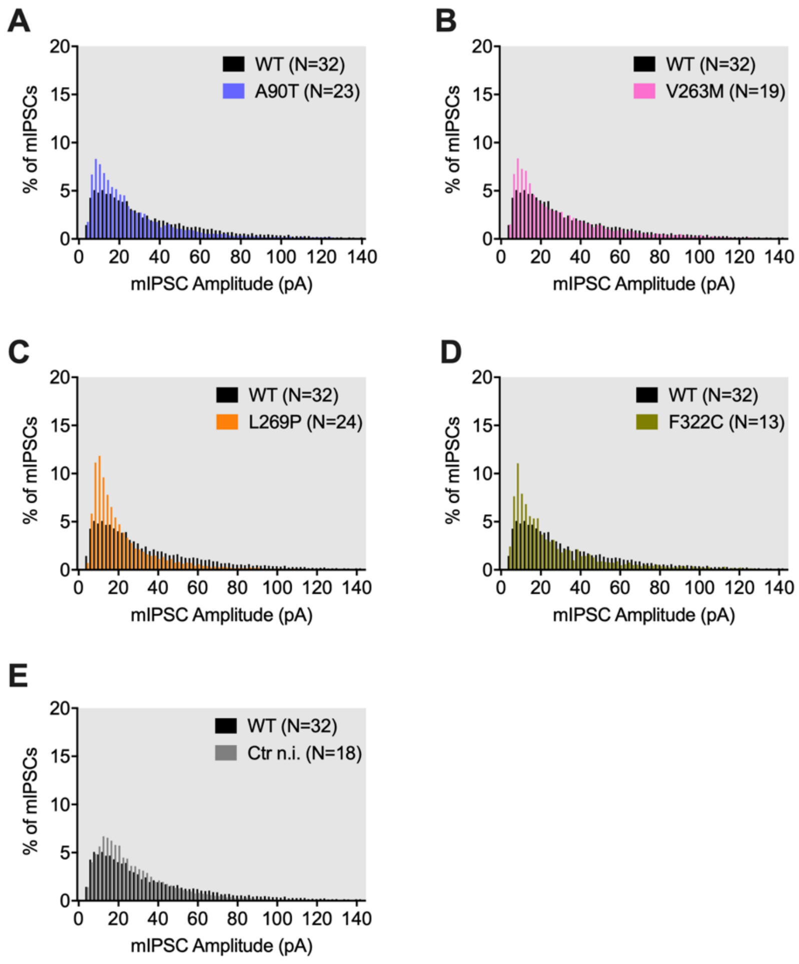

Platzer et al. Figure S4

**Figure S4:** Comparison of mIPSC amplitude distributions for *Slc32a1*$^{WT/Het}$ neurons expressing (A) VGAT$^{A90T}$ vs VGAT$^{WT}$ (B) VGAT$^{V263M}$ vs VGAT$^{WT}$ (C) VGAT$^{L269P}$ vs VGAT$^{WT}$ (D) VGAT$^{F322C}$ vs VGAT$^{WT}$. (E) Comparison of uninfected *Slc32a1*$^{WT/Het}$ neurons and neurons expressing additional VGAT$^{WT}$.
